# Tumor Patterns and Cancer Risk in Carriers of *TP53* exonic Germline Variants that alter mRNA Splicing

**DOI:** 10.1101/2025.09.22.25336167

**Authors:** Deborah Schoenegger, Emilie Montellier, Sandrine Blanchet, Claire Freycon, Paola Monti, Catherine Goudie, Gaëlle Bougeard, Christian P. Kratz, Pierre Hainaut, Anna Reymer

## Abstract

Pathogenic germline variants in the *TP53* gene cause Li-Fraumeni syndrome (LFS), a highly penetrant cancer predisposition disorder. Most of these variants arise from single-nucleotide variations (SNVs) in *TP53* exons, causing missense mutations. However, some of these SNVs may also alter mRNA splicing, defining spliceogenic single nucleotide variants (SE-SNVs) of uncertain clinical significance. We reassessed previously classified *TP53* missense variants for spliceogenic effects using SpliceAI predictions, *in vitro* minigene assays, and transcriptomic data from TCGA. Genotype-phenotype correlations were evaluated using clinical data from carriers of *TP53* germline variants across multiple databases and registries. Among 58 identified SE-SNVs, 40 were missense and 18 synonymous. Experimental validation showed that most induce aberrant splicing events, frequently via cryptic splice site activation, leading to frameshift and premature stop codons. Several missense variants previously classified as having mild or low pathogenicity were found instead to have strong spliceogenic effects and were associated with early-onset cancers typical of LFS, suggesting that splicing alterations may override their protein-coding impact. The frequent SNV c.375G>A leading to the synonymous variant p.T125= shows intermediate severity, likely due to partial retention of normal splicing activity. Our study highlights the underestimated pathogenic potential of SE-SNVs affecting the *TP53* gene. These findings underscore the importance of integrating splicing predictions, functional assays, and transcript-level analyses into *TP53* variant interpretation to improve risk stratification in LFS.

## Introduction

Abnormal splicing is a common mechanism of gene inactivation in several cancer predisposition syndromes, including the Li-Fraumeni Syndrome (LFS), a heterogeneous autosomal dominant predisposition to various cancers caused by pathogenic germline *TP53* variants^1,2^. These variants cause inactivation of the tumor suppressive properties of the p53 protein as well as, in some instances, activation of still poorly understood dominant-negative or prooncogenic effects^3,5^. The majority of LFS-related variants are single nucleotide variants (SNVs) causing missense or nonsense mutations^4^, whereas about 10% of them fall at intron/exon junctions and in intronic motifs that regulate p53 mRNA splicing. These SNVs may alter p53 mRNA splicing and p53 protein expression by inactivating splice donor or acceptor sites and/or disrupting splice enhancers/silencers. Such alterations through the usage of cryptic splice acceptors or donors, can cause partial or complete intron retention or exon skipping, often resulting in RNA nonsense—mediated decay (NMD) or, in some instances, stable but incorrectly spliced RNA that produces a functionally impaired protein^8,9^. In addition to variations at canonical splicing motifs, exonic SNVs can also modify splicing by activating otherwise cryptic splice donors/acceptors or enhancers/silencers in addition to their direct protein-coding effects^10,11^. Such spliceogenic exonic SNVs (SE-SNVs) can thus have the same structural consequences as variations at canonical splice motifs. Since they also affect codon usage, they may double-up with protein-coding alterations. The two mechanisms can thus have compounded effects, or spliceogenic effects can override those protein-coding alterations.

LFS is characterized by a lifetime risk of multiple cancers with distinct phases of cancer risk during childhood, adolescence and adulthood, defining a complex and evolving cancer spectrum that is extremely difficult to predict at individual and familial level (for detailed description of LFS tumor spectrum, see ^12,13^). Extensive clinical and functional analyses have highlighted a strong and significant correlation between variant structural characteristics, functional activity in different experimental assays and age at cancer onset as well as cancer profiles in *TP53* variant carriers^4,14–16^. In particular, we recently proposed a classification of missense variants into four functional classes, A to D, outlining a phenotypic gradient from very severe LFS (Class A) to milder forms (Classes B and C) and forms in which the characteristic LFS “signature” tumors are infrequent (Class D)^4^. However, this classification was derived from the p53 functional dataset developed by C. Ishioka and collaborators using a yeast-based transactivation assay^17^, which does not account for the potential spliceogenic effects of the SNVs underlying these missense variants.

Two recent major advances have created new opportunities for investigating the phenotypic impact of SE-SNVs. First, T. Stiewe and collaborators^11^ have developed a saturation genome editing screen with CRISPR-mediated homology-directed repair. By introducing SNVs directly in the wild-type *TP53* gene locus that includes introns and exons, this method enables for the first time a comparative analysis of the impact of any SNV on p53 functional properties in a standardized cell model, maintaining the physiological regulation of the *TP53* gene. Second, A. Spurdle and collaborators^18^ have combined bioinformatics prediction analysis and reporter minigene assays to analyze the impact on splicing of 59 potentially spliceogenic SNVs distributed across the entire *TP53* coding sequence, highlighting both the type and extent of the splicing defects they may cause and thereby providing a basis for evaluating their pathogenicity.

Building upon these recent developments, we re-assessed all the missense variants previously included in our functional classification model^4^ for the spliceogenic properties of the corresponding SNVs by combining bioinformatic predictions, *in vitro* reporter assays and transcriptomic analyses using public RNA-Seq data for from The Cancer Genome Atlas (TCGA) program. Next, we analyzed genotype–phenotype correlations in germline *TP53* variant carriers using the germline IARC/NCI database^7^ supplemented with additional curated cases from the literature (systematic review by C. Freycon, unpublished; **see Supplementary Table 1**) and French^2^ and German^6^ LFS registries, with no overlap with IARC/NCI. Our results identify several pathogenic spliceogenic SNVs generating missense variants that were initially classified as mild or benign (Classes C or D) based on their p53 functional activity changes, in which splicing effects appear to override the protein coding effects of the variant. Our results also provide the first systematic evaluation of LFS phenotypes in carriers of SE-SNVs that produce missense or nonsense mutations at the protein level, offering new insights into the molecular basis of their pathogenicity.

**Table 1:**
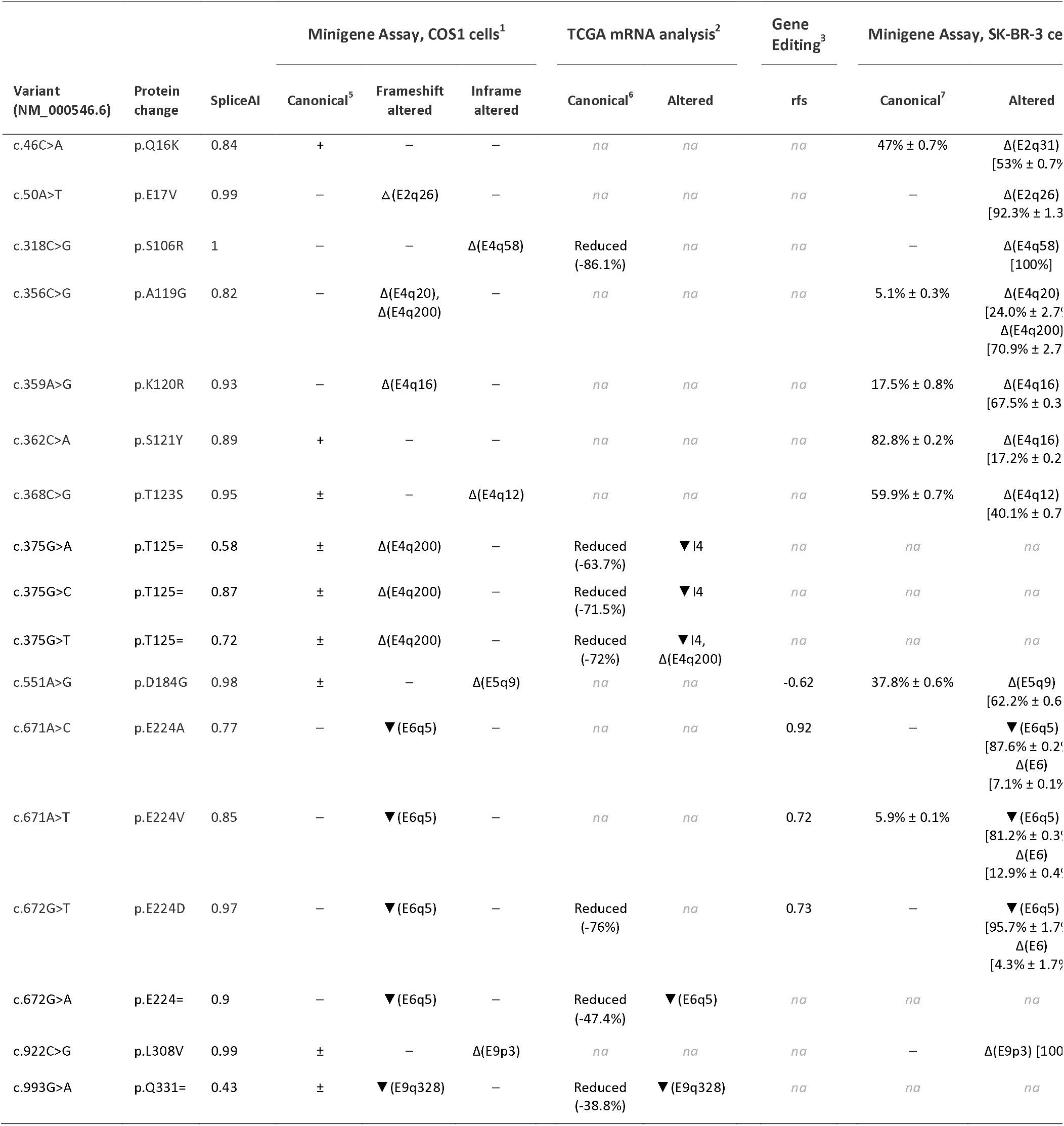
Functional and transcript-level characterization of TP53 spliceogenic exonic variants (SE-SNVs). The table lists single nucleotide variants (SNVs), corresponding protein changes, SpliceAI predictions, and observed transcript consequences, including frameshift or in-frame events, for 17 tested SE-SNVs from Class C and D with SpliceAI delta score ≥0.75 as well as those resulting in silent variants with SpliceAI delta scores ≥ 0.4. Presented are data generated in the current study: minigene assays in COS1 cells (1) and TCGA mRNA analysis (2); together with published functional results from CRISPR/Cas9 gene editing^11^ (3) and minigene assays in SK-BR-3 cells^18^ (4). Levels of normally spliced p53 mRNA (“canonical”) were estimated by semi-quantitative sizing of sequencing peaks (5; stratified as +, ±, -), mRNA read counts (6; % of total reads), or semi-quantitative PCR (7; % of total p53 mRNA). Symbols: Δ = exon skipping (or partial skipping); *▼* = intron retention (or partial retention). q/p notation indicates nucleotide position within the exon (q = downstream portion) or intron (p = upstream portion) relative to the splice junction. NA = not available.

## Results

### SpliceAI Predictions of *TP53* SNVs

We first analyzed the predicted impact on splicing of SNVs in the *TP53* coding sequence (exons 2 to 11) using SpliceAI, a deep neural network that predicts splice junctions from any arbitrary pre-mRNA transcript sequence, enabling the identification of noncoding as well as coding SNVs that may cause splicing defects^19^. We selected as exonic spliceogenic SNVs (SE-SNVs) single nucleotide variants with SpliceAI delta scores ≥ 0.4, a threshold that combines high sensitivity (97.9%) with reasonable specificity (72.4%) (AUC: 0.95; **Supplementary Figure 1**). We found that candidate SE-SNVs, which might otherwise result in missense or silent variants, are predicted across the entire *TP53* coding sequence, with distinct density patterns observed across different exons (**Figure 1A**). A total of 58 SE-SNVs were identified (**Supplementary Table 2**). Of these 58 SNVs, 18 resulted in synonymous (silent) variants and 40 resulted in missense variants. A total of 42 of these SNVs were documented in either germline or somatic *TP53* databases, forming a small proportion of these datasets (COSMIC: 40/1761, 0.02%; IARC/NCI *TP53* somatic database: 58/3409, 0.02%, *TP53* germline database: 13/314, 0.04%; gnomAD: 12/730, 0.02%). The most represented SE-SNVs in these datasets generated synonymous substitutions at codons 125 (c.375G>T, c.375G>A, c.375G>C; p.T125=), 224 (c.672G>C; p.E224=), and 331 (c.993G>A; p.Q331=), as well as missense substitutions at codons 106 (c.318C>G; p.S106R), 119 (c.356C>G; p.A119G), 120 (c.359A>G; p.K120R), 187 (c.559G>A; p.G187S and c.559G>C; p.G187R), 224 (c.672G>T, c.672G>C; p.E224D), and 331 (c.993G>T, c.993G>C ; p.Q331H). A comparison between somatic and germline datasets showed that the same SNVs tended to occur in both datasets, suggesting that the alterations they cause can contribute to carcinogenesis in both contexts (**Figure 1A, Supplementary Table 2**). However, their distribution appeared to be more restricted in the germline as compared to the somatic dataset. In the germline dataset, the single SE-SNV, c.375G>A (p.T125=) accounted for 38% of the occurrences (14 families, 59 cases), followed by c.559G>A (p.G187S; 11%), c.375G>T (p.T125=; 11%), c.375G>C (p.T125T=; 8%) and c.993G>A (p.Q331=; 7%), with no other variant accounting for more than 3% of the dataset. In contrast, in the somatic (COSMIC) dataset, the most frequent SE-SNVs were c.375G>T (p.T125=; 17%), followed by c.375G>A (p.T125=; 13%), c.559G>A (p.G187S, 8%), c.672G>T (p.E224D, 7%), c.318C>G (p.S106R, 6%), c.672G>A (p.E224=, 6%), c.993G>T (p.Q331H; 5%) and remaining variants representing less than 5% of the dataset (**Supplementary Table 2**).

**Figure 1:**
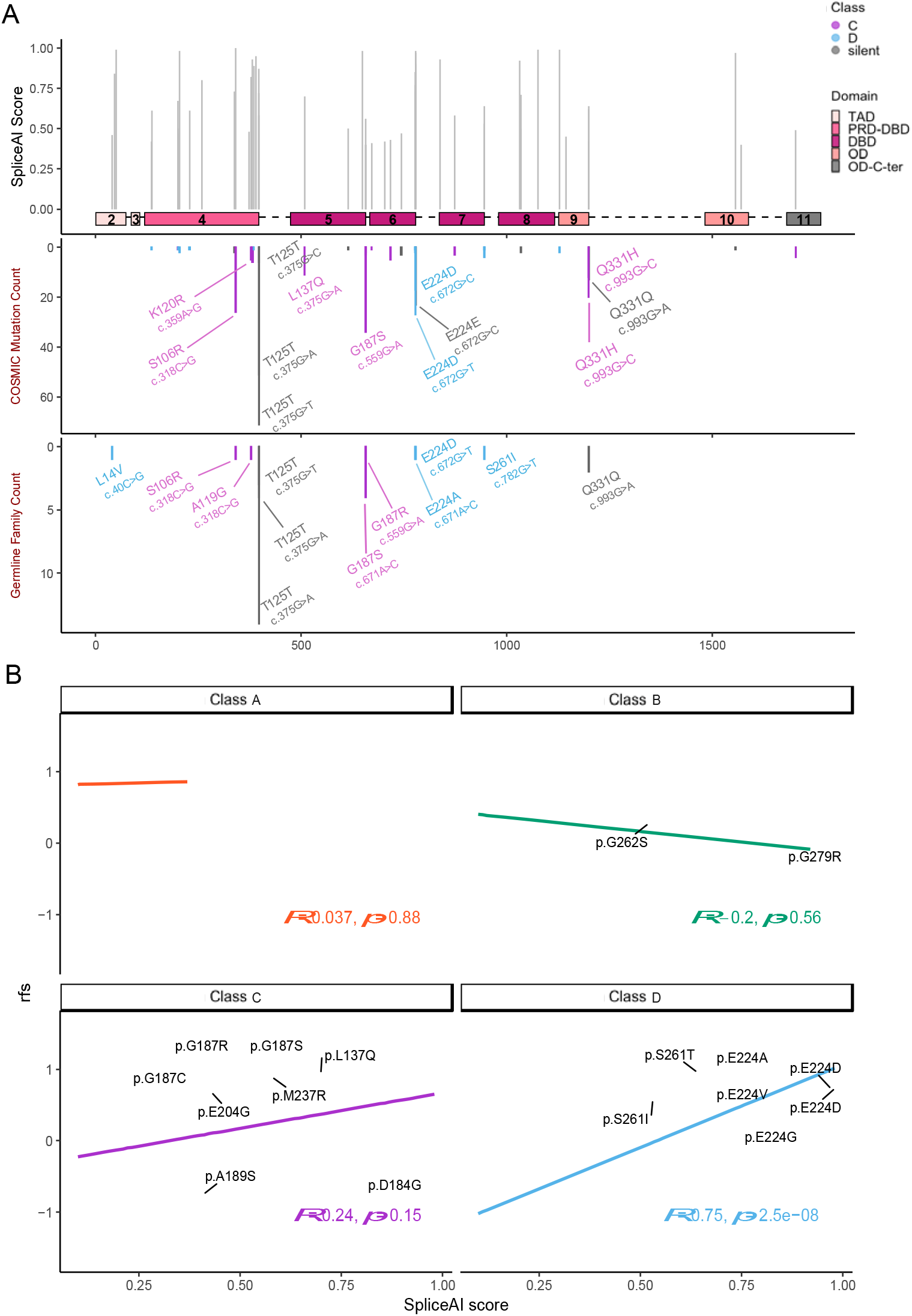
Distribution of Spliceogenic Exonic Single Nucleotide Variations (SE-SNVs) in the *TP53* gene and relationships with missense variant functional classes. A: Top panel: SpliceAI delta scores versus location of SE-SNVs (Class C: blue; D: violet; synonymous: grey) variants along the *TP53* coding sequence (threshold: SpliceAI ≥0.4, see text). Exons are represented as colored boxes with their corresponding numbers, introns (not to size) are outlined as dotted lines. Lower panels: Distribution of SE-SNVs among COSMIC (middle panel) and germline (bottom panel) *TP53* datasets (see text). B: Correlation between the relative fitness score (rfs)^11^, and SpliceAI delta scores of SE-SNVs, stratified by *TP53* functional class^4^. Red: Class A (very severe variants, including *TP53* hotspot variants, recapitulating all severe traits of LFS); Green: Class B (severe variants); Blue: Class C (heterogeneous variants associated with less severe LFS traits); Violet: Class D (mostly predicted benign variants not associated with typical LFS cancer spectrum). Rfs ranges from -1 (functional, wild-type reference) to +1 (non-functional, similar to nonsense mutations. Variants with SpliceAI delta scores ≥ 0.4 are labeled by their protein change.

Missense variants were annotated according to the functional classes A to D proposed by P. Hainaut and collaborators^4^, outlining a gradient of loss of transcriptional activity in a standardized in *vitro* assay that predicts a phenotypic gradient of severity in germline *TP53* variant carriers (from Class A: most severe variants, matching the criteria of Pathogenic/Likely Pathogenic variants, to Class D: variants retaining quasi-wild-type p53 activities, matching functional definitions of Benign/Likely Benign variants). Candidate SE-SNVs were predominantly identified among Classes C and D, as well as among silent (synonymous) variants (91.4%, **Figure 1A, Supplementary Table 2**), whereas only 8.6% of predicted SE-SNVs belonged to Classes A or B (**Supplementary Table 2**).

To further evaluate the functional impact of these SE-SNVs according to their corresponding missense variant class, we analyzed the correlation between SpliceAI delta scores and the relative fitness scores in the CRISPR-mediated homology-directed repair assay developed by T. Stiewe and collaborators^11^ (**Figure 1B, Supplementary Table 3**). The fitness scores (rfs) vary from -1 (*i.e.*, variants retaining a quasi-wild-type activity) to +1 (*i.e.*, the most severely impaired variants). As expected, Class A and B SE-SNVs tended to have higher rfs values than Class C or D variants, consistent with their more severe structural and functional alterations. For these classes, SpliceAI delta scores showed no correlation with rfs. In contrast, among Class C and especially Class D variants, a positive correlation between the two scores was observed, reaching strong statistical significance in Class D (p < 0.001). Overall, these findings suggest that some Class C and D variants, classified as attenuated or benign based solely on the functional impact of their amino acid substitutions, may exert pathogenic effects through splicing alterations that override their protein-coding consequences.

### Functional Analysis of SE-SNVs

We next verified the SpliceAI-predicted splicing defects using a pSPL3-based *in vitro* minigene assay in COS1 fibroblast-like monkey cells. The vector contains two endogenous exons, separated by a cloning site for inserting short genomic DNA segments carrying the putative SE-SNVs. The *TP53* genomic sequence was divided in three segments: the first segment comprising exons 2-4, the second – exons 5–8, and the third – exons 8-10 (**Supplementary Figure 2**). SE-SNVs within each of these segments were tested for their ability to produce an altered pattern of splicing as compared with the control vector (i.e., empty pSPL3) containing the two endogenous exons V1 and V2 separated by a standard intron; an example of the redouts of this assay are shown in the lower panel of **Supplementary Figure 2**. We used this assay to determine the effects on splicing of Class C and D SE-SNVs with SpliceAI delta score values ≥ 0.75, as well as those resulting in silent variants with SpliceAI delta scores ≥ 0.4. Among 17 tested SE-SNV, 15 (88%) produced altered splicing patterns caused by the inserted variant splice motifs and resulting in either skipping of exonic nucleotides or inclusion of intronic nucleotides (**Table 1, Supplementary Figure 3**). In most cases, these events predicted frameshift and premature stop codons, causing nonsense-mediated RNA decay. Importantly, for several variants, these altered splicing events co-existed with the normal splicing pattern of wild-type p53. Only two variants scored as negative in this assay, c.46C>A (p.Q16K) and c.362C>A (p.S121Y), with sequencing revealing only normal splicing events and no evidence of cryptic splice site activation. A particular splicing pattern was observed for c.551A>G (p.D184G), for which the minigene assay revealed an in-frame deletion causing the loss of three amino-acid residues (D184-S185-D186) at the end of exon 5, as well as partial retention of normal splicing patterns. Of note, this SNV with the corresponding p.D184G variant appeared as an outlier among Class C in **Figure 1B**, with a SpliceAI delta score of 1 (predicting high spliceogenicity) but the low rfs score (-0.62, predicting quasi-wild-type p53 functionality). Whether the resulting p53 protein with the in-frame deletion retains at least partially wild-type activity is not known.

To gain further insight into how these SE-SNVs affect splicing patterns *in vivo*, we analyzed public RNA-Seq transcriptomic data from The Cancer Genome Atlas (TCGA). Raw RNA-Seq read alignments were examined to determine the exact structure of SE-SNV–associated splicing events and to compare them with those observed in the minigene assay. Of the 17 SE-SNVs, seven (41.2%) have been identified in solid tumors in the public TCGA dataset. Analysis of these cases confirmed strong spliceogenic effects, consistent with the results from the minigene assay (**Table 1, Supplementary Figure 4**). In the case of the c.375G>T SNV, which results in the synonymous variant p.T125=, RNA-Seq reads revealed intron retention in addition to the cryptic splice site activation observed in the minigene system.

To further characterize the effects of these SE-SNVs, we compared our minigene assay and RNA-Seq read alignment results with those of Fortuno et al.^18^, who used a conceptually similar minigene assay incorporating *TP53* exons 2–9 but based on a different cell model (i.e., SK-BR-3 breast cancer cells versus non-transformed COS1 cells in our assay) (**Table 1**). The comparison revealed high concordance between the two datasets. Notably, the SK-BR-3–based assay detected additional events not observed in our COS1-based assay, including: (1) for c.671A>C (p.E224A), c.671A>T (p.E224V), and c.672G>T (p.E224D), exon 6 skipping in addition to the retention of five intronic nucleotides also detected by us; and (2) for c.46C>A (p.Q16K) and c.362C>A (p.S121Y), skipping of 31 nucleotides in exon 2 (ΔE2q31) and skipping of 16 nucleotides in exon 4 (ΔE4q16), respectively, whereas these SNVs showed only canonical splicing patterns in our analyses.

### Genotype-Phenotype Correlations

We next analyzed cancer onset and tumor phenotypes among germline carriers of SE-SNVs identified by SpliceAI, minigene assays, and transcriptomic analyses, and compared them with carriers of other protein-altering variants. Specifically, we aimed to determine whether carriers of SE-SNV previously classified as Class C or D variants are associated with more severe phenotypes than predicted by their protein-coding effects. To do so, we assembled an extended dataset of carriers with SE-SNVs (synonymous or Class C/D variants) by integrating three sources: (i) the public IARC/NCI germline database^7^ (version 20, last updated 2025), (ii) additional cases curated in a recent systematic literature review (2019–2025; C. Freycon, unpublished; **see Supplementary Table 1**), and (iii) non-redundant cases from the French^2^ and German^6^ LFS clinical registries. This analysis identified 18 carriers of SE-SNVs classified as Class C (c.318C>G; p.S106R, c.356C>G; p.A119G, c.559G>C; p.G187R, c.559G>A; p.G187S) or Class D (c.672G>T; p.E224D, c.40C>G; p.L14V, c.671A>C; p.E224A, c.782G>T; p.S261I) variants, who collectively developed 23 cancers (diagnosis age range: 2–50 years). For comparison, we examined: (1) carriers of non–SE-SNVs Class A, B, C, or D variants (n = 1426, 290, 238, and 171, respectively); (2) carriers of SNVs at intronic splice sites (n = 219), included as a reference group of splice-disrupting mutations, since they are either located at canonical splice sites or annotated as experimentally validated to alter splicing in the IARC/NCI database; and (3) carriers of the SE-SNV c.375G>A resulting in a synonymous variant p.T125=, the most frequent variant in the germline *TP53* dataset (n = 59 carriers who developed 61 cancers) (**see Supplementary Table 4**).

Results (**Figure 2A–D**) showed that carriers of Class C and D SE-SNVs had a median age at cancer diagnosis of 27 years, similar to that of carriers of Class A variants (28 years) and comparable to carriers of intronic splice variants (30 years). In contrast, carriers of non–SE-SNV Class C or D variants had a markedly higher median age at diagnosis (≥40 years). These findings suggest that, for Class C or D SE-SNVs, spliceogenic effects may override the impact of the amino-acid substitution. Notably, p.T125= *TP53* variant carriers had a median diagnosis age of 38 years, intermediate between that of Class B and Class C variant carriers (**Figure 2D**). The distribution showed a bimodal pattern (**Figure 2D**), consistent with a variable severity profile: a substantial fraction of cases occurred in childhood and adolescence, while others presented later in adulthood, underscoring that this variant can manifest across a wide severity spectrum. The number of carriers (n) for each SNV category analyzed in this study is provided in **Supplementary Table 4**.

**Figure 2:**
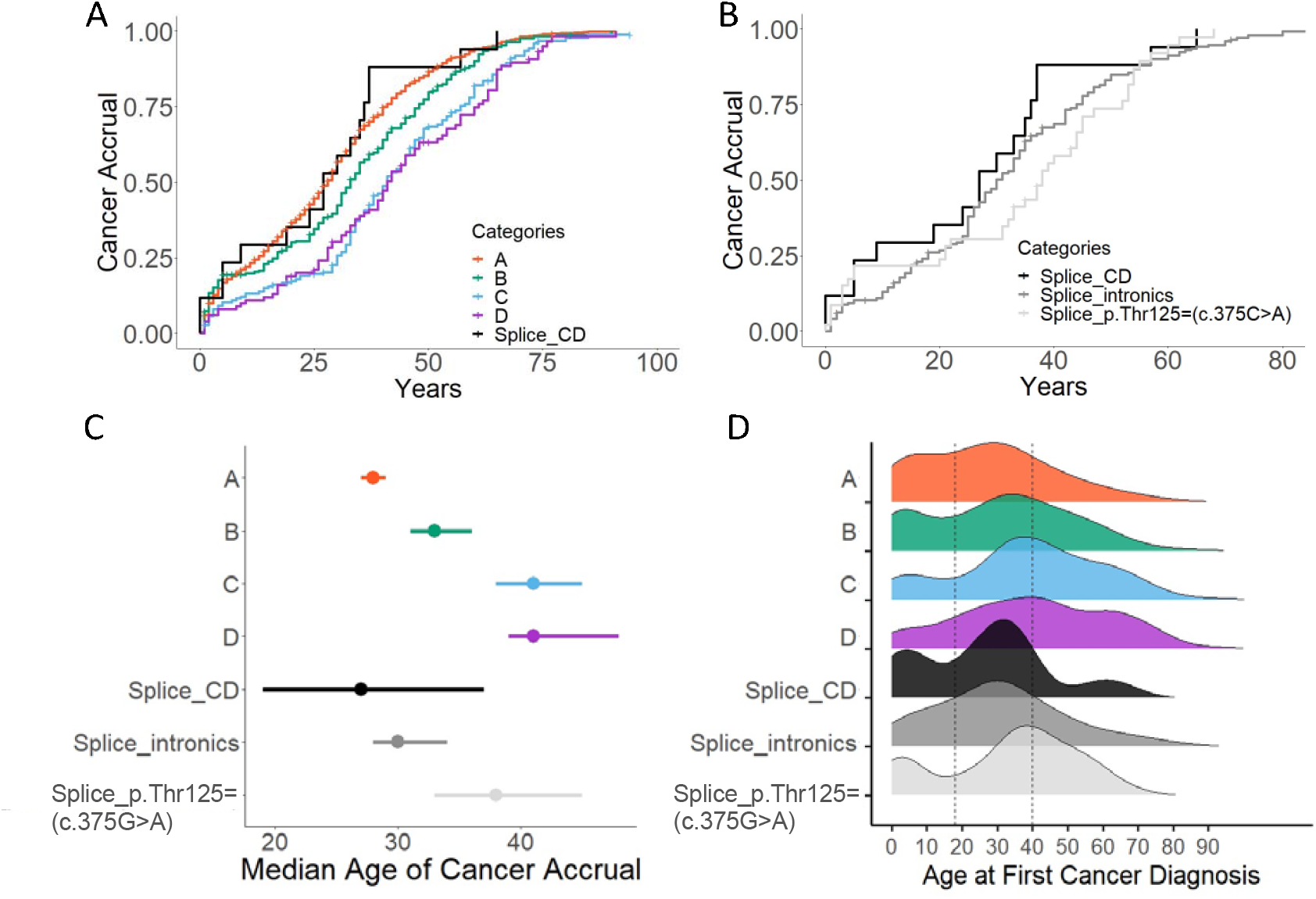
Genotype-phenotype correlations for SE-SNVs among carriers of germline *TP53* variants. A: Cancer accrual in carriers of SE-SNVs, previously classified as Class C-D variants (black line) compared to intronic spliceogenic *TP53* SNVs and non-SE deriving missense variants stratified by functional Class A–D, color-coded as in Figure 1. B: Cancer accrual in carriers of SE-SNVs previously classified as Class C-D variants (black line) compared with carriers of intronic SE-SNVs (canonical splice donor/acceptor sites, dark grey line) or of the synonymous SE-SNV c.275G>A (p.T125=; light grey line). C: Density plots showing the age distribution of the first cancer diagnosis for each variant category, as defined and color-coded above. D: Median age at first cancer diagnosis and 95% confidence intervals for each variant group, color coded as above.

The age distribution of first cancers (**Figure 2C**) and of all cancers (**Supplementary Figure 5**) further supported that Class C or D SE-SNVs confer a more severe cancer risk than their non-spliceogenic counterparts, with a greater proportion of cases arising in childhood and adolescence. Tumor types in these carriers included childhood adrenal cortical carcinoma, teenage osteosarcoma and astrocytoma, and pre-menopausal breast cancer: all characteristic of the LFS tumor spectrum (**Supplementary Figure 6**). Notably, the age distribution showed two distinct peaks: one in childhood/teenage years and another in early adulthood. Similarly, carriers of the *TP53* p.T125= (c.375G>A) variant exhibited tumor types typical of the LFS spectrum, but with onset shifted toward adulthood compared with Class A or B carriers. Among the three documented carriers of the *TP53* p.Q331= (c.993G>A) variant, one developed adrenal cortical carcinoma between ages 5–10, while two developed breast cancer between ages 35– 40. Overall, these findings support the hypothesis that SE-SNVs can produce a more severe cancer phenotype than would be predicted from the amino-acid substitution alone. However, the limited sample size, particularly for Class C and D SE-SNV carriers, precluded robust statistical confirmation.

Together, the 18 carriers of Class C/D SE-SNVs identified here account for 4.8% of all Class C and 3.4% of all Class D germline *TP53* variants in our dataset (**Supplementary Table 4**). These proportions give an indication of their relative frequency in the LFS setting but should be interpreted with caution, given dataset incompleteness, differences in ascertainment, and the inclusion of additional cases from systematic review and the national registries.

## Discussion

In this study, we combined bioinformatic predictions, experimental analysis using a minigene reporter assay, and transcriptomic analysis of *TP53* RNA splicing in tumors to assess the spliceogenic effects of single nucleotide variants occurring in the coding exons (SE-SNVs) of the gene. We confirm and further demonstrate that several SE-SNVs, which might otherwise result in synonymous or missense changes, also have significant spliceogenic effects, causing a broad range of disruptions, including partial or complete exon/intron skipping, frameshifts, introduction of premature stop codons, and nonsense-mediated mRNA decay. Next, we identified a subset of such SE-SNVs in *TP53* germline variant carriers and show that the severity of the cancer phenotype aligned more closely with the predicted spliceogenic effects of the single nucleotide *TP53* variant than with the predicted impact of the amino acid substitution. This effect was particularly strong in carriers of SE-SNVs previously assigned to Classes C or D based on their transcriptional activity in a yeast transcriptional activation assay^4,17^. These carriers showed a more severe phenotype than carriers of non-spliceogenic SNVs encoding Class C or D variants, recapitulating all the characteristic features of the LFS tumor spectrum. Their tumor patterns and age at cancer onset were similar to those of carriers of intronic splice variants, a group representing well-established splice-disrupting mutations, and only marginally less severe than those of carriers of pathogenic Class A or B missense variants. These observations suggest that the spliceogenic effects overrode those of their amino acid substitutions, supporting their reclassification as severe variants based on the effect of the nucleotide change rather than the amino acid change, consistent with Pathogenic/Likely Pathogenic ClinVar annotations^14^. However, these genotype–phenotype correlation analyses were constrained by the small size of the SE-SNV carrier dataset and the intrinsic heterogeneity of LFS phenotypes, which together preclude robust statistical evaluation. In addition, the analyzed cohorts were ascertained phenotypically, whereas more accurate risk estimates could be achieved through the analysis of genomically ascertained cohorts.

Our genotype-phenotype correlations also highlight the heterogeneity and diversity of the phenotypic effects of SE-SNVs. In particular, we show that carriers of c.375G>A (p.T125=), the most frequent SE-SNV associated with LFS, displayed variable cancer onset and phenotypes compared to carriers of Class A or B variants or of intronic splice variants. Of note, c.375G>A occurs within a methylated CpG site, a mutation-prone position due to spontaneous deamination of 5-methylcytosine to thymine^20^. This unique feature among codons specifying SE-SNVs may explain its frequent occurrence among LFS patients. The phenotypic variability may reflect incomplete disruption of normal splicing, as suggested by detection of a low level of normally spliced transcripts in our assays. However, such findings from minigene and tumor RNA-Seq analyses cannot fully capture physiological complexity. Overall, these results are consistent with the notion that the impact of SE-SNVs depends not only on their ability to induce aberrant splicing and nonsense-mediated mRNA decay but also on the degree of residual normal splicing, which may vary between tissues and influence pathogenicity.

The recent study by Fortuno et al.^18^ systematically explored the role of splicing in *TP53* variant pathogenicity using SpliceAI predictions and minigene assays. These analyses provide a robust basis for evaluating and comparing the predicted functional impact of different types of spliceogenic variants. The authors concluded that SpliceAI predictions generally aligned well with minigene results, but that the maximum SpliceAI delta score did not accurately predict the level of aberrant expression. Our results confirm and reinforce this conclusion. SpliceAI, a deep neural network trained on genome-wide human transcriptome annotations from the GENCODE and GTEx projects, predicts spliceogenic effects by modeling long-range sequence dependencies (up to 10,000 bp) around each position^19,21^. This algorithm captures both core splice site signals and auxiliary splicing regulatory features, indirectly integrating biological determinants such as RNA secondary structure, chromatin accessibility, and nucleosome positioning that influence splice site recognition.

In our analyses, a SpliceAI delta score ≥ 0.4 correctly identified all SE-SNVs (resulting in synonymous and missense changes) annotated in either germline (IARC/NCI) or somatic (COSMIC) *TP53* mutation databases. However, SpliceAI also assigned high scores (≥ 0.8) to several variants not yet documented in these cancer databases. Among these, SE-SNVs encoding Class D variants such as c.551A>G (p.D184G), c.46C>A (p.Q16K), and c.368C>G (p.T123S) showed at least partial retention of normal splicing patterns (40–60%) in our assay. While these findings suggest that residual splicing may suffice to maintain some p53 functionality, quantitative estimates from minigene systems should be interpreted with caution, as they do not fully recapitulate endogenous splicing and require validation in patient-derived samples. In contrast, other SE-SNVs resulting in Class D variants, such as c.50A>T (p.E17V) and c.671A>C (p.E224A), showed near-complete absence of wild-type splicing patterns in minigene assays and are thus probably pathogenic. In the case of c.671A>C (p.E224A), its CRISPR-mediated introduction into the *TP53* locus caused near-complete loss of p53 suppressive activity (rfs = 0.92^11^). Consistent with this predicted severity, this variant was recently identified in a family fulfilling Chompret’s criteria for LFS^22^.

Despite the overall concordance, our minigene assay results showed differences from those of Fortuno et al.^18^, who detected additional aberrant splicing events beyond those we observed. This was particularly evident for SE-SNVs at codon 224 (p.E224A, p.E224V, p.E224D), for which Fortuno et al.^18^ reported exon 6 skipping in addition to the retention of five intronic nucleotides also detected in our assay, and for SE-SNVs resulting in p.Q16K and p.S121Y, which showed altered splicing in their study but only canonical splicing in ours. One key methodological difference is that our assay used non-transformed COS1 cells (a fibroblast-like kidney cell line from the African green monkey; ATCC CRL 1650), whereas Fortuno et al.^18^ used the human breast cancer cell line SK-BR-3. While SK-BR-3 cells are not known to carry mutations directly affecting the splicing machinery, splicing dysregulation is a hallmark of most cancer types, leading to both alternative and atypical splicing events. Such relaxed splicing control in cancer cells may therefore explain the additional events observed in their assay. This phenomenon might also account for the differing representation of SE-SNVs in somatic versus germline mutation datasets, as certain nucleotide variants may preferentially, or even exclusively, induce splicing defects in the somatic context of cancer cells. In addition, splicing outcomes are inherently variable and often tissue-specific, which is a critical consideration when evaluating phenotypic consequences in Li-Fraumeni syndrome. A further experimental difference is that Fortuno et al.^18^ employed a single construct spanning *TP53* exons 2–9, whereas we used three separate constructs covering exons 2–4, exons 5–8, and exons 8–10. Additional comparative studies will be valuable to determine how different experimental designs influence splicing readouts and to ensure accurate classification of *TP53* variants in germline carriers.

In conclusion, our results underscore that *TP53* missense variants previously classified as benign or likely benign based on protein structural and functional properties can, in fact, cause profound splicing defects leading to severe LFS phenotypes. Although SE-SNVs producing such missense variants are rare in LFS patients and families, they should not be overlooked, particularly in individuals with borderline or atypical LFS presentations. While SpliceAI predictions offer a reliable first step in detecting potential splice defects, our findings emphasize the need for in-depth functional characterization to assess variant pathogenicity. Such evaluation should include both the identification of aberrantly spliced transcripts and the quantification of residual correctly spliced p53 mRNA, as this is not accurately predicted by SpliceAI and may influence pathogenicity depending on the functional integrity of the encoded p53 protein. Implementing allele-specific transcript analysis in peripheral blood cells, providing both qualitative and quantitative profiles of transcripts from each allele, would allow more precise classification of *TP53* variants and ultimately improve risk assessment and clinical management for LFS patients.

## Materials and Methods

### Distribution and Frequencies of *TP53* Missense Mutations with Predicted Effect on Splicing

We retrieved all possible single nucleotide substitutions in *TP53* exonic sequences from the IARC/NCI *TP53* Database (R21 version, https://tp53.isb-cgc.org/). SpliceAI (v.1.3)^23,24^ was used for *in-silico* prediction of splicing effects. The SpliceAI delta score, defined as the maximum predicted probability that a variant alters splicing at any position within a ±10 kb window (recommended for optimal performance), was recorded for each substitution. A delta score threshold of ≥ 0.4 was applied, as this value provided a balance between sensitivity (97.9%) and specificity (72.4%) in receiver operating characteristic (ROC) analysis (AUC = 0.95) using a publicly available validation webtool (https://gwiggins.shinyapps.io/lr_shiny/, standard settings) developed by S. Topper and collaborators^25^, with threshold selection consistent with approaches described in Wu et al.^26^

Frequencies of missense *TP53* mutations with a SpliceAI delta score ≥ 0.4 have been analyzed in three different datasets: 1) cancer–related germline *TP53* variants (IARC/NCI *TP53* Database, https://tp53.isb-cgc.org/; R21 version), 2) cancer–related somatic variants (COSMIC database; downloaded on November 12, 2024; https://cancer.sanger.ac.uk/) and 3) non-cancer related germline variants (gnomAD, v. 4.1.0, https://gnomad.broadinstitute.org/).

### Functional Impact of Predicted SE-SNVs

Missense variants were assigned to functional Classes A–D according to the classification proposed by P. Hainaut and collaborators^4^, which exploited the yeast-based transactivation assay dataset developed by developed C. Ishioka and collaborators^17^. We also integrated data from a CRISPR–Cas9–mediated homology-directed repair saturation mutagenesis screen^11^, which assessed the proliferative fitness of *TP53* variants in the DNA-binding domain (exons 5–8) following p53 pathway activation by MDM2 inhibition. Enrichment scores from this assay were normalized to relative fitness scores (rfs), ranging from −1 (synonymous mutations) to +1 (nonsense mutations). For each variant, rfs values were correlated with the corresponding SpliceAI delta scores using Pearson regression (ggplot2 and ggpubr R packages, version 2023.12.1+402).

### Minigene Splicing Assay

We used the exon trapping vector pSPL3^27^ (Burn et al., 1995) according to the modified protocol of Tompson and Young^28^, with simian green monkey kidney COS1 cells as the recipient cell line. The pSPL3 vector contains a small artificial gene composed of an SV40 promoter, an exon–intron–exon sequence with functional splice donor and acceptor sites, and a late polyadenylation signal. The single intron includes a multiple cloning site that can accommodate a genomic fragment to create a minigene expression construct. We divided the *TP53* cDNA into three fragments (“boxes”): Box 1: exons 2–5 (1,701 nt), Box 2: exons 5–8 (1,678 nt), and Box 3: exons 8–10 (3,142 nt); each fragment was flanked by 50 intronic nucleotides upstream and downstream of the canonical donor/acceptor splice sites (**Supplementary Figure 2**). SE-SNVs with SpliceAI delta scores ≥ 0.75 and located within 60 nt of a splice junction were introduced into these constructs by site-directed mutagenesis. Additional constructs were generated for the synonymous variants p.T125= (c.375G>A, c.375G>T, c.375G>C), p.E224= (c.672G>A), and p.Q331= (c.993G>A).

All constructs were synthesized and sequence-verified by Azenta Life Sciences – Genewiz (Leipzig, Germany). Plasmids were transformed and amplified in *E. coli* TOP10 (Thermo Fisher, Ref C505003) and purified for transfection. COS1 cells were seeded at 4 × 10^5^ cells/well in 6-well plates and transfected at 70–80% confluency according to the Tompson and Young^28^ protocol, with minor modifications. Total RNA was isolated using the NucleoSpin RNA Plus kit (Macherey-Nagel, Düren, Germany; Ref 740990.50). Reverse transcription was performed with the iScript™ Reverse Transcription Supermix kit (Bio-Rad, Providence, RI; Ref 1708840). PCR amplification was carried out using primers V1-F (5⍰-TCTGAGTCACCTGGACAACC-3⍰, exon 1) and V2-R (5⍰-ATCTCAGTGGTATTTGTGAGC-3⍰, exon 2) with the Platinum SuperFi DNA Polymerase kit (Thermo Fisher, Ref 12351010). PCR products were resolved by agarose gel electrophoresis, purified, and analyzed by Sanger sequencing (GENEWIZ from Azenta Life Sciences, Leipzig, Germany).

### TCGA RNA-Seq Analysis

For SE-SNVs tested in the minigene assay, corresponding mRNA expression data were retrieved, when available, from the TCGA RNA-Seq datasets (https://www.cancer.gov/ccg/access-data) and compared with expression levels of wild-type *TP53*. RNA-Seq level-3 data from 33 cancer cohorts were downloaded from Firebrowse (http://firebrowse.org), and raw read counts were normalized across the combined dataset using the DESeq2 package (v1.46.0), excluding genes with fewer than 10 total read counts across all samples. mRNA expression levels for each patient were matched to their mutation profiles obtained from cBioPortal (https://www.cbioportal.org/). Differences in *TP53* expression between mutant and wild-type samples were calculated as the percentage change (reduction or increase) relative to wild-type *TP53* expression levels. To examine splicing patterns, raw RNA-Seq data (NCI dbGaP Study Accession: phs000178.v11.p8) were analyzed, and splice junction usage was visualized using Sashimi plots generated in R with the ggsashimi tool^29^.

### *TP53* Germline Variant Dataset

Clinical information for carriers of Class C and D SE-SNVs was compiled from the germline *TP53* variants database (https://tp53.cancer.gov/), an extended literature search, and Li–Fraumeni syndrome registries in France and Germany^2,6^. Carriers of missense variants from Class A-D, as well as carriers of the splice variant c.375G>A (p.T125=), were identified from the germline *TP53* variants database. Individuals harboring multiple *TP53* variants were excluded because the contribution of each variant could not be reliably determined. Carriers of the Brazilian founder variant *TP53* p.R337H who were not recruited through a family history of cancer were also excluded to avoid analytical bias.

### Genotype–Phenotype Correlations

Lifetime cancer incidence by variant class was assessed using either the age at first cancer diagnosis or the cancer-free age of individuals. Median age at cancer onset and 95% confidence intervals (CIs) were calculated. The age distribution of all cancers, including both primary and secondary malignancies, was analyzed. Tumor distribution within each class was determined based on topography, with organ groups relevant to Li–Fraumeni syndrome (adrenal gland, brain, bone, soft tissue, breast, hematopoietic system, and other organs) consolidated for analysis. All analyses were performed in R.

### Nomenclature

Variants are described according to HGVS recommendations; for synonymous variants we use HGVS-compliant notation (e.g., p.T125=).

## Data Availability

All data produced in the present study are either publicly available or contained within the manuscript and supplementary files. Public datasets were obtained from the IARC/NCI TP53 Database, COSMIC, gnomAD, and TCGA (NCI dbGaP Study Accession: phs000178.v11.p8). Data generated from minigene assays are available within the manuscript and supplementary materials.

## Dedication

The authors dedicate this work to the memory of Prof. Pierre Hainaut, whose scientific vision and mentorship continue to guide and inspire us.

## Acknowledgements

The results shown here are in part based upon data generated by the TCGA Research Network: https://www.cancer.gov/tcga.

The authors thank Dr. Renata Bocciardi (IRCCS Istituto Giannina Gaslini, Genoa, Italy) for her valuable advice regarding the splicing vector.

Studies on *TP53* variants at IAB Grenoble were supported by the ERiCAN program of the Fondation MSD Avenir (to P.H. and E.M.). A.R. was supported by an Inserm Research Chair (*Inserm Chaire de Recherche*). C.P.K. was supported by the BMBF ADDRess program (01GM2205A) and by the Deutsche Kinderkrebsstiftung (DKS 2024.03). P.M. was supported by E.T.S. Engineering Ship Technology Pte Ltd.

