## Supplementary Information for "Tumor Patterns and Cancer Risk in Carriers of *TP53* exonic Germline Variants that alter mRNA Splicing"

[†] Deceased

### Table of Content:

**SI Table 1**. Sources of SE-SNV carriers included in the genotype–phenotype analyses, compiled from IARC/NCI, systematic literature review, and LFS registries. — p. 2

**SI Figure 1**. Receiver Operating Characteristic (ROC) curve evaluating SpliceAI performance in predicting splice-disruptive variants. — p. 3

**SI Tables 2 and 3**: attached as a separate Excel file.

**SI Figure 2**. *TP53* minigene constructs expressed in COS-1 cells, including schematic representation and RT-PCR analysis. — p. 4

**SI Figure 3**. Sanger sequencing electropherograms of minigene assay results with indications of cryptic splice site activation. — p. 5-6

**SI Figure 4**. Sashimi plots showing raw RNA-Seq read alignments from TCGA patients carrying selected *TP53* SE-SNVs (p.S106R, p.T125=, p.Q331=) alongside normal tissue controls. — p. 7-9

**SI Figure 5**. Density plots showing the age distribution of all cancer diagnoses for each variant category. — p. 10

**SI Figure 6**. Distribution of cancer types among carriers of missense variants (Class A–D), spliceogenic Class C/D variants, intronic splice variants, and the synonymous mutation p.T125=. — p. 11

**SI Table 4**. Number of carriers for each category analyzed in this study, and the corresponding proportions. — p. 12

**References.** — p. 13

**SI Table 1:** Sources of SE-SNV carriers Included in the genotype–phenotype analyses.

| Database/Source | SE-SNV | ProtDescription | YTA_class[3] | Reference |
|---|---|---|---|---|
| NCI/IARC Germline | c.318C>G | p.S106R | C | PMID: 11518751 |
| NCI/IARC Germline | c.672G>T | p.E224D | D | PMID: 19930417 |
| NCI/IARC Germline | c.356C>G | p.A119G | C | PMID: 30076369 |
| Literature | c.40C>G | p.L14V | D | PMID: 26189108 |
| Literature | c.559G>C | p.G187R | C | PMID: 30239254 |
| Literature | c.559G>A | p.G187S | C | PMID: 34771502 and 39962599 |
| LFS registries | c.559G>A | p.G187S | C | LFS registry (France)[4] |
| LFS registries | c.671A>C | p.E224A | D | LFS registry (France)[4] |
| LFS registries | c.559G>C | p.S261I | D | LFS registry (France)[4] |
| LFS registries | c.356C>G | p.A119G | C | LFS registry (Germany)[5] |

The table documents the provenance of the SE-SNV cases analyzed in this study. Cases were compiled from three sources: (i) the IARC/NCI germline *TP53* database[6] (version 20), (ii) a systematic literature review conducted between 2019–2025, and (iii) the French[4] and German[5] Li-Fraumeni syndrome (LFS) clinical registries. Duplicate entries across datasets were removed. This compilation is provided to ensure transparency of case selection and traceability of references.

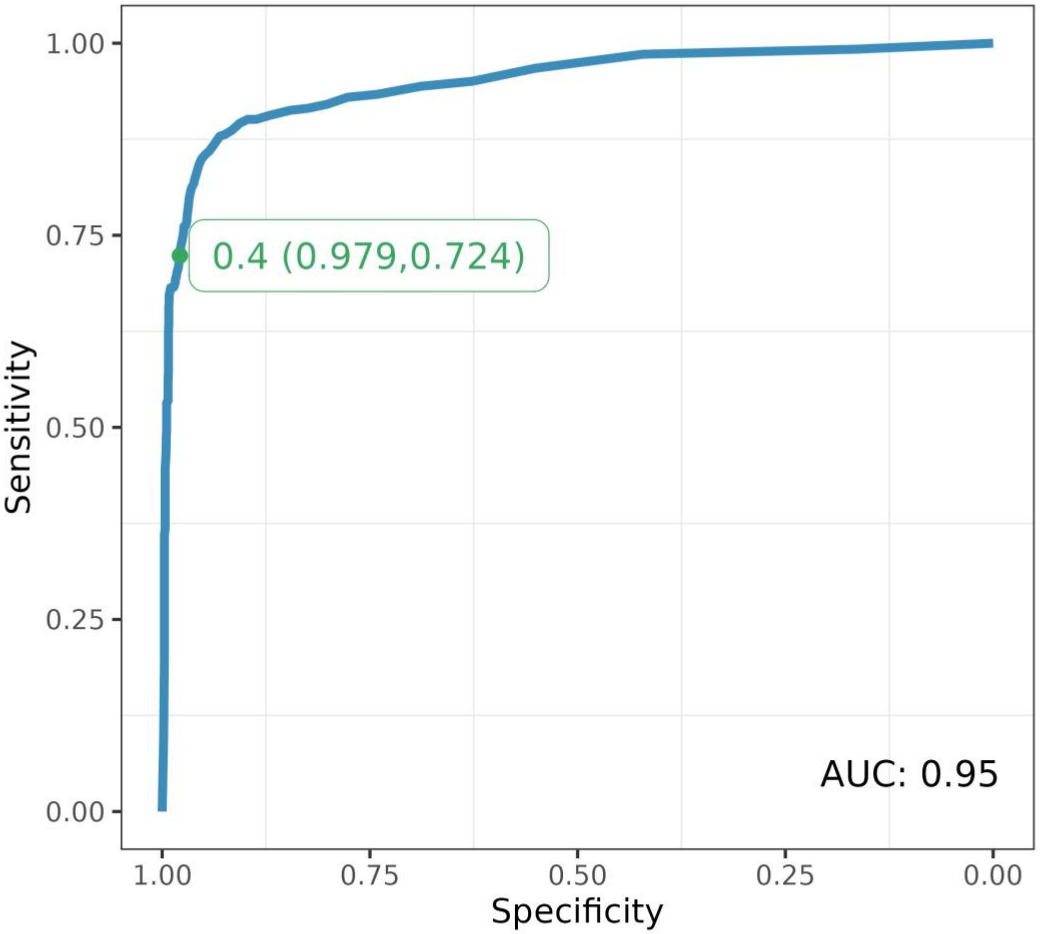


**SI Figure 1**: Receiver Operating Characteristic (ROC) curve evaluating SpliceAI performance in predicting splice-disruptive variants. The curve illustrates the trade-off between sensitivity and specificity across varying SpliceAI score thresholds. A threshold of 0.4 is highlighted, yielding a sensitivity of 0.979 and a specificity of 0.724. The area under the curve (AUC) is 0.95, indicating strong predictive performance. This analysis is based on a curated dataset of 3,021 variants across BRCA1/2, mismatch repair genes (*MLH1, MSH2, MSH6, PMS2*), *NF1*, and *POU1F1*, which includes *in vitro* splicing assay results[1].

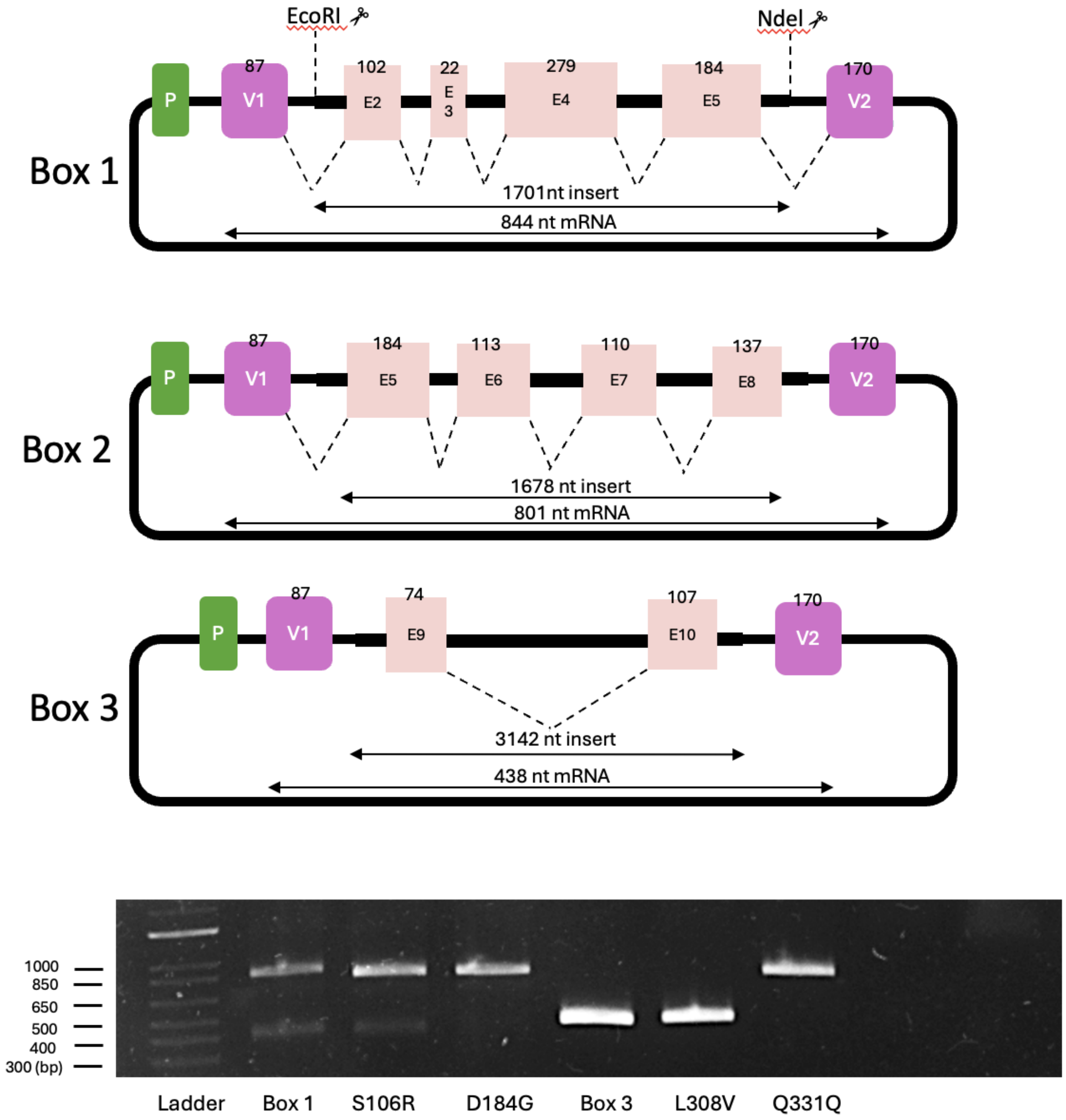


**SI Figure 2**: *TP53* minigene constructs expressed in COS-1 cells. Genomic regions of the *TP53* gene were divided into three overlapping fragments ("Box 1," "Box 2," and "Box 3") and cloned into the pSPL3 exon trapping vector, which contains vector-specific exons V1 and V2 and a promoter (P). Each construct contains selected *TP53* exons (E2–E10) and flanking intronic sequences, indicated by exon numbers and dashed lines. Arrows indicate the expected size of the inserted genomic DNA (nt insert) and the spliced mRNA (nt mRNA) following expression in COS-1 cells.

The lower panel shows an example of RT-PCR analysis on RNA extracted from COS-1 cells transfected with wild-type (Box1 and Box3) or mutant minigene constructs. PCR products were visualized in agarose gel and PCR products were Sanger sequenced to verify alternative splicing patterns.

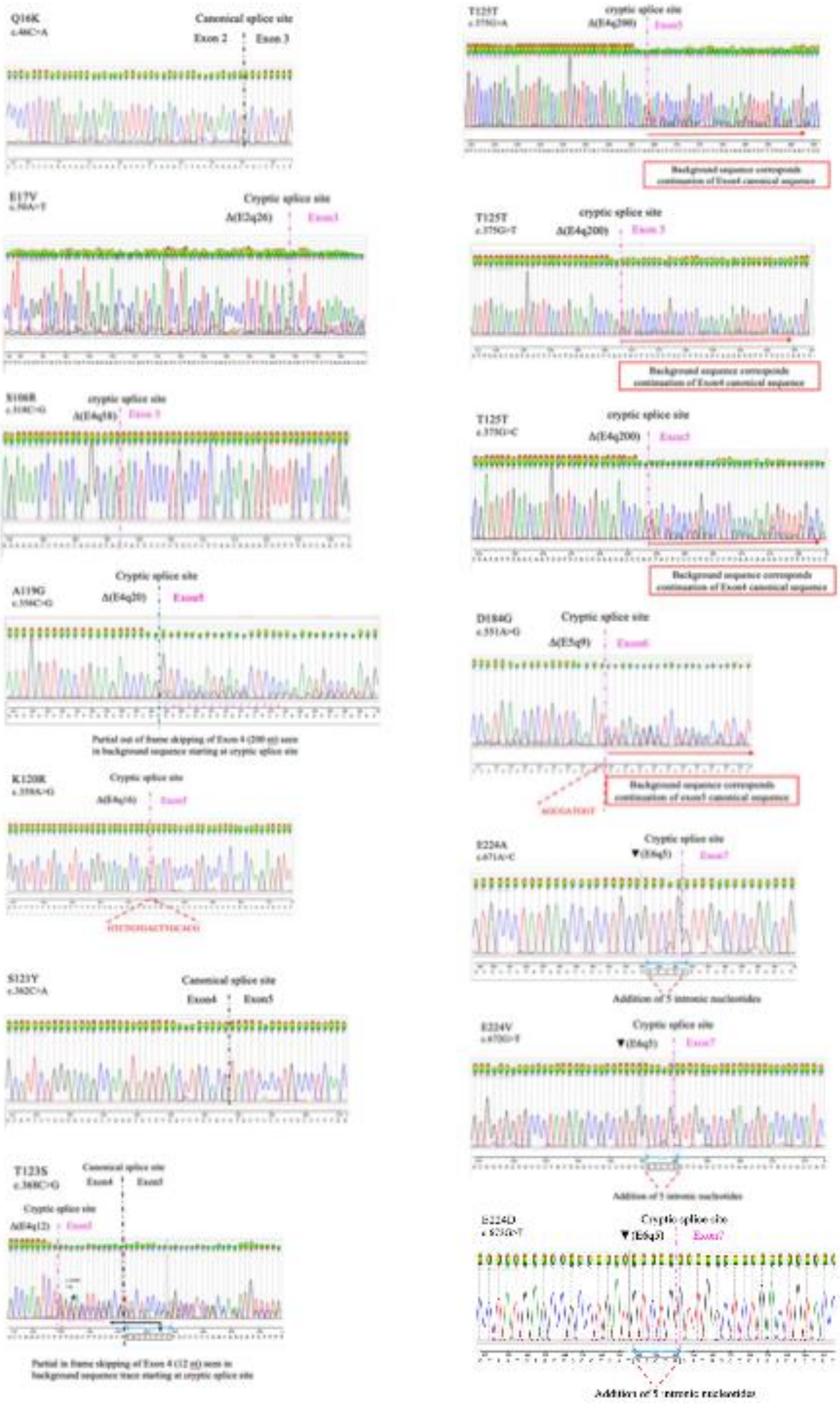

Q16K
Canonical splice site
Exon 2
Exon 3
E17V
Cryptic splice site
S106R
cryptic splice site
A119G
Cryptic splice site
Δ(E4q20)
Partial out of frame skipping of Exon 4 (200 nt) seen in background sequence starting at cryptic splice site
K120R
Cryptic splice site
S121Y
Canonical splice site
Exon4
Exon5
T123S
Canonical splice site
Cryptic splice site
Partial in frame skipping of Exon 4 (12 nt) seen in background sequence trace starting at cryptic splice site
T125T
cryptic splice site
Δ(E4q200)
Background sequence corresponds continuation of Exon4 canonical sequence
T125T
cryptic splice site
Δ(E4q200)
Exon 5
Background sequence corresponds continuation of Exon4 canonical sequence
T125T
cryptic splice site
Δ(E4q200)
Background sequence corresponds continuation of Exon4 canonical sequence
D184G
Cryptic splice site
Δ(E5q9)
Background sequence corresponds continuation of exon5 canonical sequence
E224A
Cryptic splice site
Addition of 5 intronic nucleotides
E224V
Cryptic splice site
Addition of 5 intronic nucleotides
E224D
Cryptic splice site
Exon7
Addition of 5 intronic nucleotides

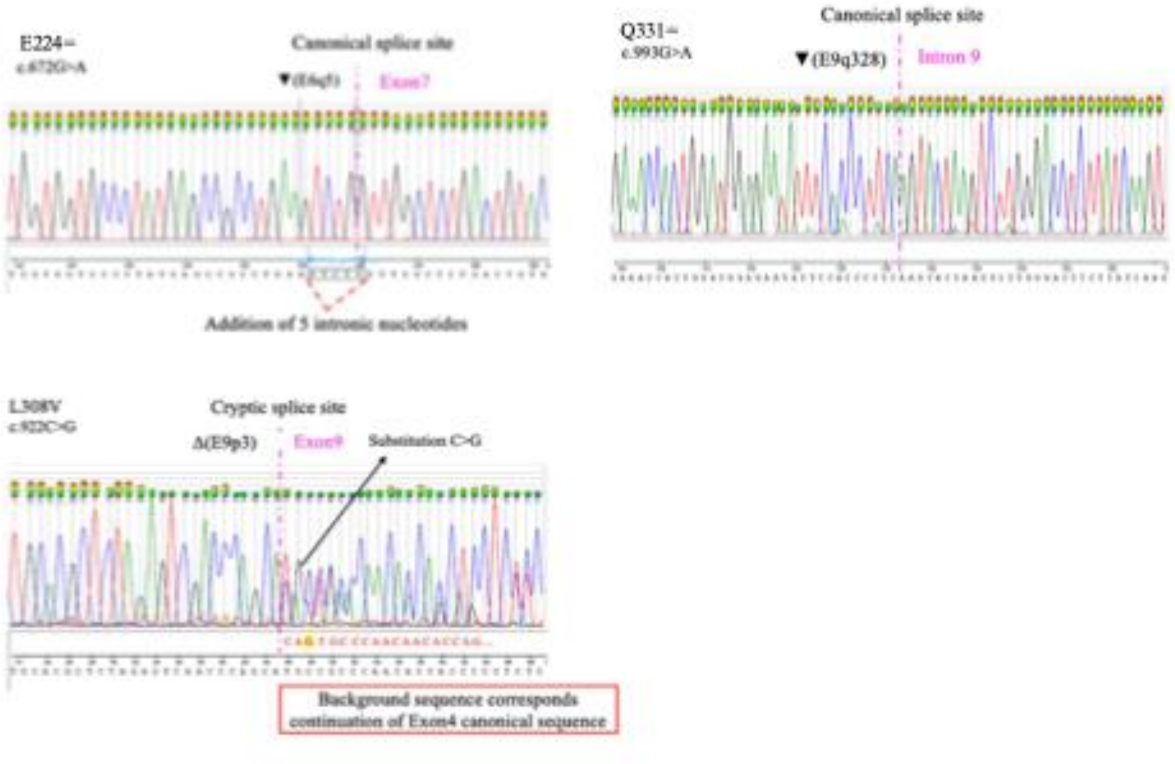


**SI Figure 3**: Sanger sequencing electropherograms of minigene assay results with indications of locations of cryptic splice sites.

A

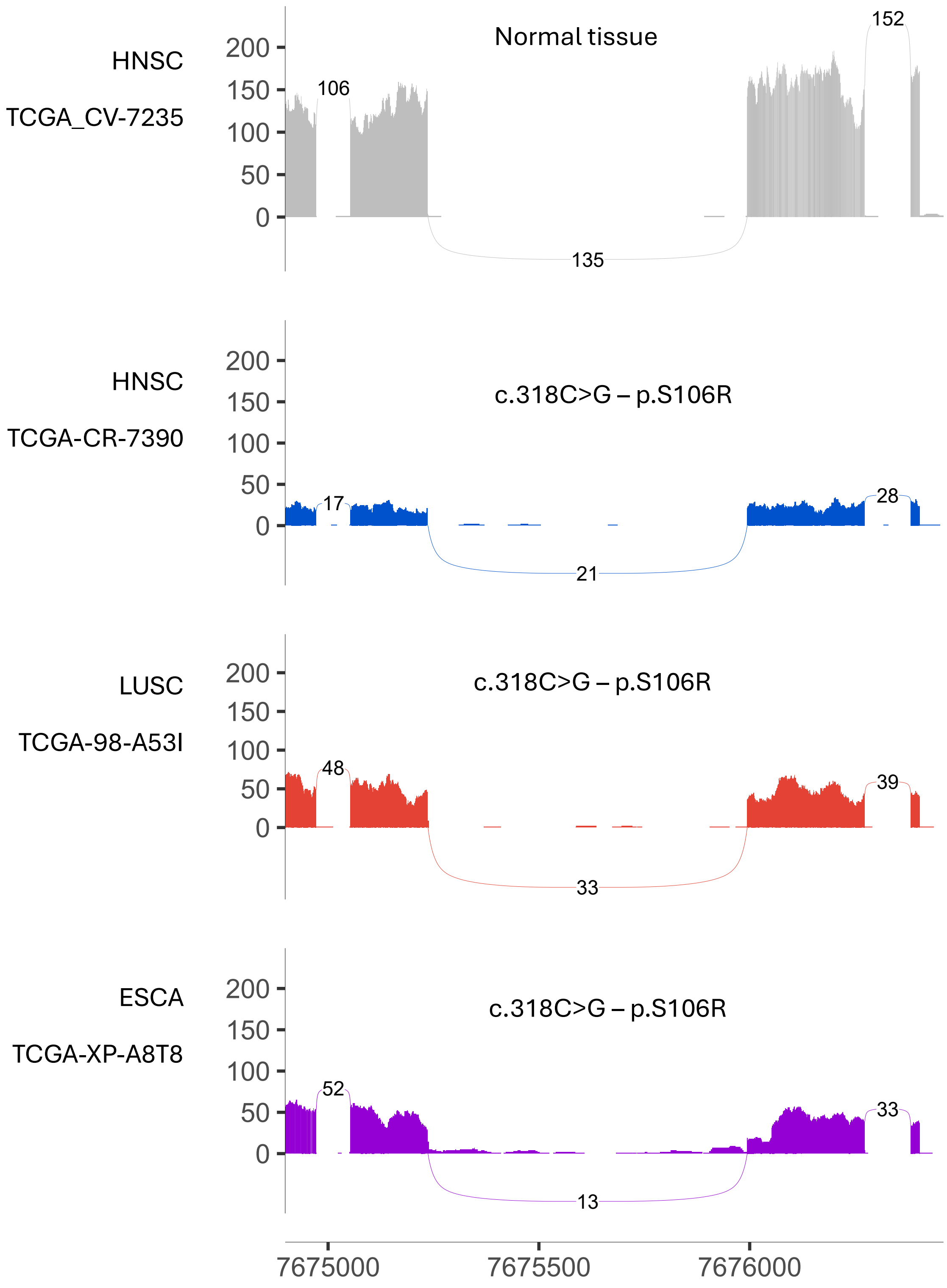

HNSC
TCGA_CV-7235
Normal tissue
200
150
100
50
0
106
152
135
HNSC
TCGA-CR-7390
c.318C>G – p.S106R
17
28
21
LUSC
TCGA-98-A53I
c.318C>G – p.S106R
48
39
33
ESCA
TCGA-XP-A8T8
c.318C>G – p.S106R
52
33
13
7675000
7675500
7676000

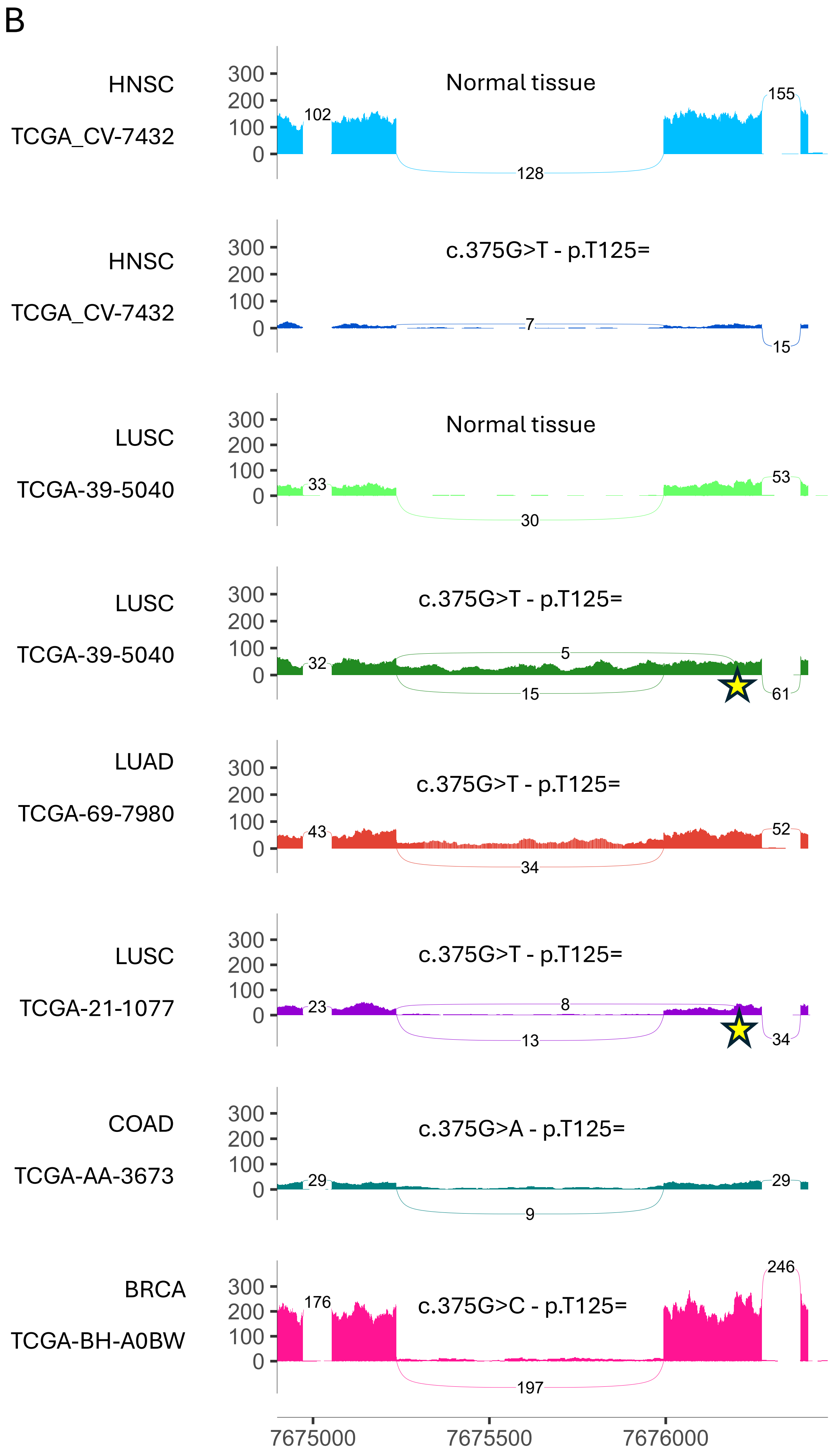
B
HNSC
TCGA_CV-7432
Normal tissue
102
155
128
HNSC
TCGA_CV-7432
c.375G>T - p.T125=
7
15
LUSC
TCGA-39-5040
Normal tissue
33
53
30
LUSC
TCGA-39-5040
c.375G>T - p.T125=
32
5
15
61
LUAD
TCGA-69-7980
c.375G>T - p.T125=
43
52
34
LUSC
TCGA-21-1077
c.375G>T - p.T125=
23
8
13
34
COAD
TCGA-AA-3673
c.375G>A - p.T125=
29
29
9
BRCA
TCGA-BH-A0BW
c.375G>C - p.T125=
176
246
197
300
200
100
0
7675000
7675500
7676000

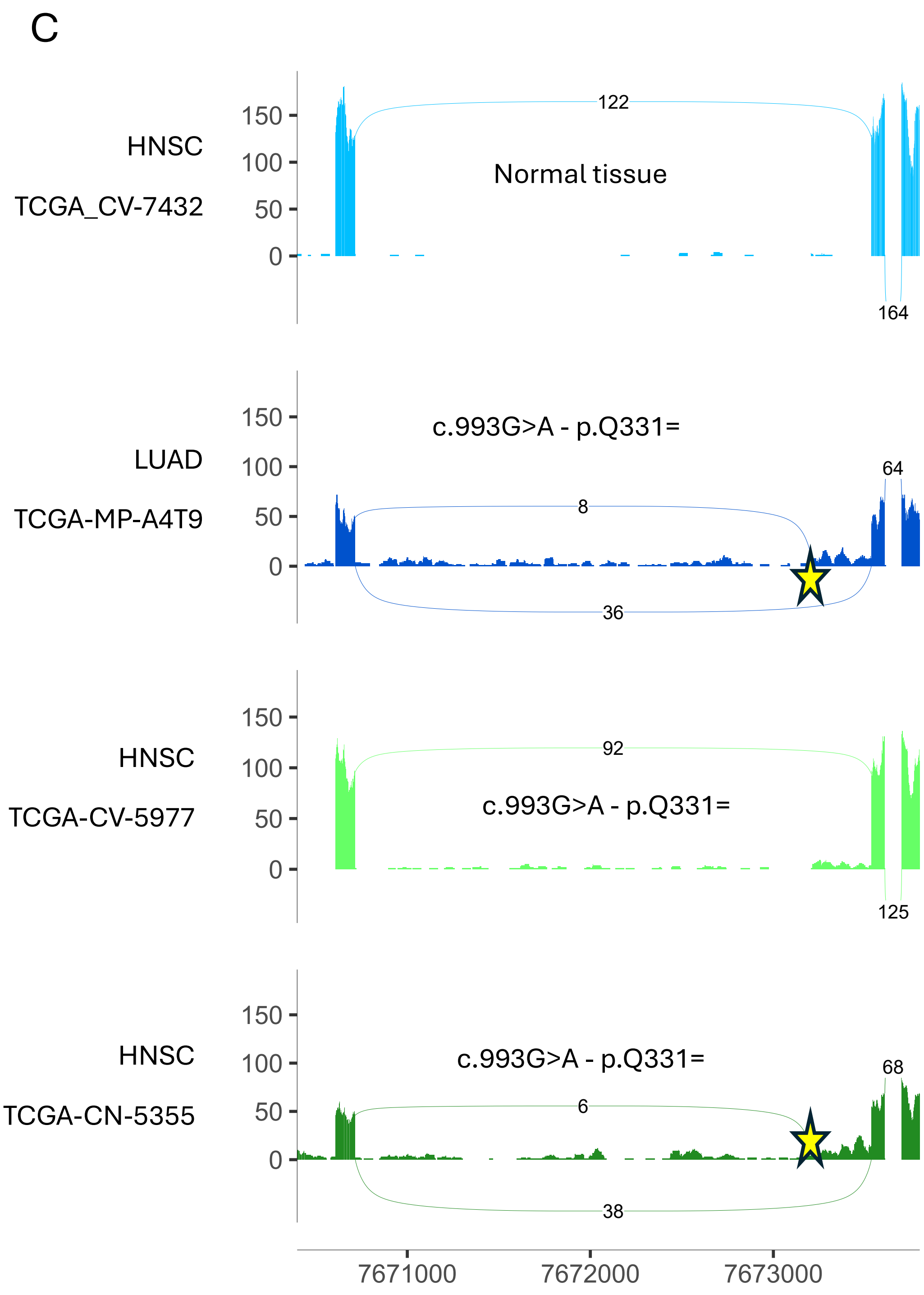


**SI Figure 4**: Sashimi plots showing raw read alignments from TCGA patients with **A**: p.S106R *TP53* variant (c.318C>G); **B**: T125= (c.375G>T, c.375G>A and c.375G>C); and **C:** Q331= (c.993G>A) mutations together with a normal tissue sample in each case as comparison. Sashimi plots of variants at codon 224, namely E224D (c.672G>C or c.672G>T) and E224= (c.672G>A) are shown in Velkova et al.[2] LUSC, lung squamous cell carcinoma; LUAD, lung adenocarcinoma; HNSC, head and neck squamous cell carcinoma; COAD, colon adenocarcinoma; ESCA, esophageal carcinoma; BRCA, breast cancer.

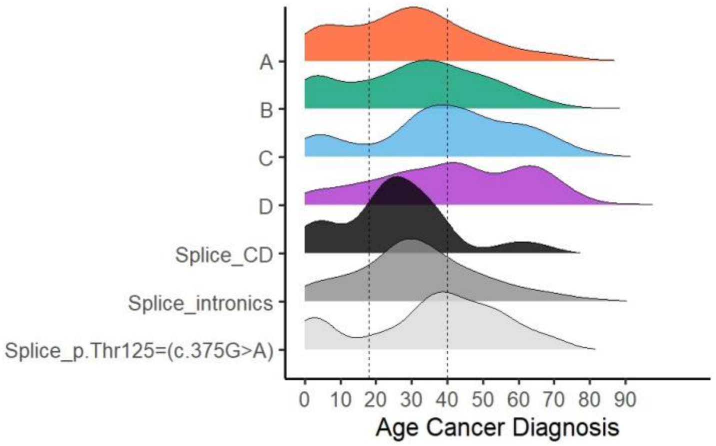


**SI Figure 5**: Density plots showing the age distribution of all cancer diagnoses for each variant category.

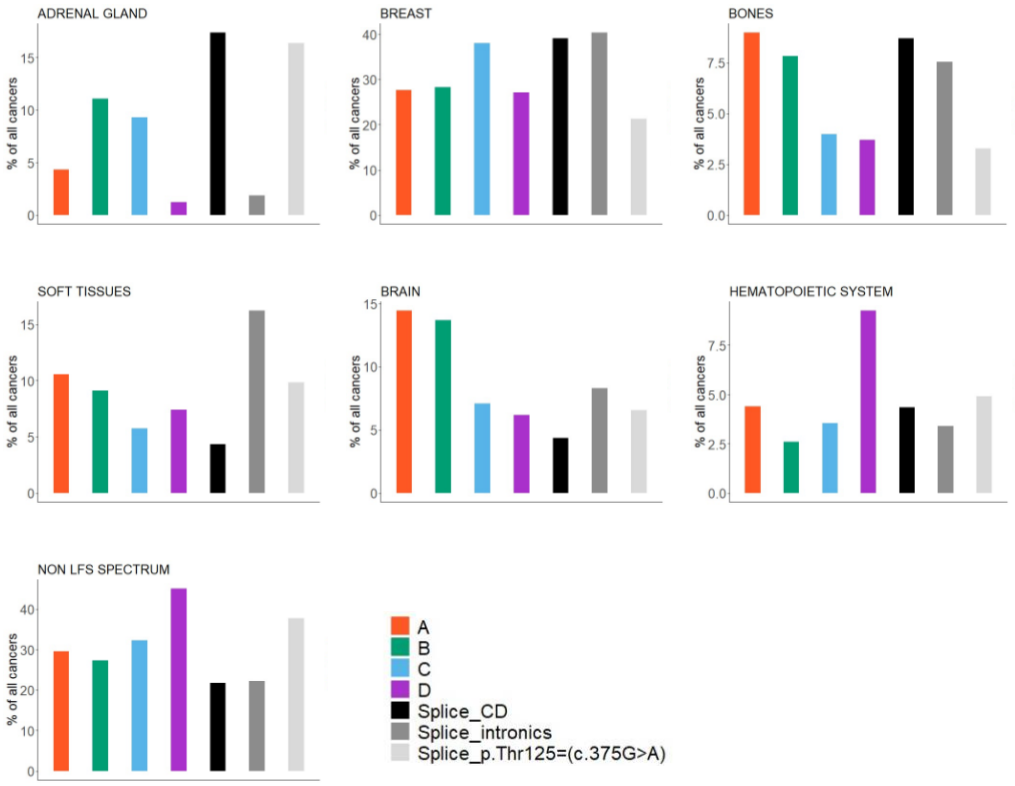


**SI Figure 6**: Distribution of cancer types for missense *TP53* variants of class A-D, class C and D missense mutations with predicted spliceogenic effects (25 cancers in total), intronic splice mutations and the synonymous mutation T125= (c.375G>A).

**SI Table 4:** Number of carriers for each category analysed in the current study, and the corresponding proportions.

| Category/Class | Number of individuals | Proportion |
|---|---|---|
| A | 1426 | 59% |
| B | 290 | 12% |
| C | 238 | 10% |
| D | 171 | 7% |
| Splice_CD* (Splice_C + Splice_D) | 18 (12 + 6) | 1% |
| Splice_intronics | 219 | 9% |
| Splice_p.Thr125=(c.375G>A) | 59 | 2% |
| TOTAL | 2421 | 100% |

*Proportion of CD_splice variants relative to the total number of individuals in Class C and D *TP53* variants according to Montellier et al.[3]:

C = 238 | **Splice_C** = 12 (**4.8% of class C** individuals)

D = 171 | **Splice_D** = 6 (**3.4% of class D** individuals)

Total C+D = 409 | Total **Splice_CD** = 18 (**4.2% of class C+D**)
